# The Australian Congenital Heart Disease Registry - Linkage to National Administrative Health Data

**DOI:** 10.64898/2026.02.04.26345613

**Authors:** Calum Nicholson, Geoff Strange, Larissa Lloyd, William Baxter, David S. Celermajer, the Congenital Heart Alliance of Australia and New Zealand InvesQgators

## Abstract

**Background:** Congenital Heart Disease (CHD) research must focus on outcomes that affect the whole-of-life course. To achieve this, datasets with long term follow up and patient-relevant outcomes are required. This paper reports on the linkage of The Australian and New Zealand Congenital Heart Disease Registry (ANZCHD Registry) (>80 000 unique individuals) with Australian National Administrative Health records and describes the final dataset.

**Methods:** Linkage on two cohorts was conducted by accredited linkage agencies, after all appropriate Ethics and Governance approvals. Cohort 1 included people who were identified from the ANZCHD Registry and Cohort 2 included people with an inpatient admission with a CHD diagnosis who had not been identified in Cohort 1. Healthcare events linked from 2010 to 2024 included outpatient encounters and medications, hospital admissions and emergency department presentations. Linked data was cleaned and curated to minimize the impacts of errors from the probabilistic linkage process.

**Results:** The final dataset included 94,383 subjects with structural CHD (58,523 from Cohort 1 and 35,860 from Cohort 2). There were over 35 million linked healthcare events recorded for this population, from 2010 to 2025. Cohort 1 was younger by an average of 14 years (95% CI: 13.2 – 13.9, p<0.001) and had a higher proportion of severe CHD lesions (20%) compared to Cohort 2 (6%) (χ2 = 7433.1, p<0.001).

**Conclusions:** The linkage described here represent a significant enrichment of the large and comprehensive Australian National CHD Registry. This will provide important research infrastructure that will enable better quality research in CHD.

**Key Messages:**

- We sought to link the Australia and New Zealand Congenital Heart Disease Registry with comprehensive, national Australian administrative healthcare records.
- The final dataset included a total of 95,383 individuals with over 35 million healthcare events from 2010 to 2025.
- Congenital Heart Disease is a whole-of-life condition with a growing and ageing population and comprehensive datasets such as these need to be made available to improve healthcare for people with Congenital Heart Disease.

## Introduction

Congenital Heart Disease (CHD) is a “whole-of-life” condition where considerable investment in pediatric care has greatly increased overall life expectancy (1-3), creating a need for increased investment in adult care (4). The changing epidemiology of the CHD population (5) has highlighted the need for research to shift its focus to long-term outcomes and the nature of the life course of the disease (6). Creating datasets that are suitable for the analysis of long-term outcomes is challenging. Whilst CHD is the most common birth-defect (7), children and adults with CHD present as a very heterogeneous population where specific lesions are quite rare. Data with long follow up periods are necessary to account for this variability and to provide the statistical power required to describe the life course of CHD accurately.

Several groups have sought to progress CHD Research from smaller single center studies to multi-center and national scale datasets, developing longitudinal CHD Registries aimed at enabling detailed research in CHD at a population level (8-11). These disease-specific registries provide high quality data relevant for the specialist area, often with the specified coding required to adequately detail the nature and complexity of the CHD. Administrative record linkage of electronic health data (for example, linking with datasets that show use of medications; hospital presentations; outpatient visit; vital status) is another valuable methodology, to optimize understanding of outcomes and service utilization, for example. With these datasets often providing population level data with potentially decades of follow up, researchers are demonstrating the benefits of using such data for epidemiology and population health (12, 13).

Indeed, several groups have begun to implement these techniques for CHD populations, linking outcomes to genetic variation (14), exploring mortality and survival (15, 16) and describing patterns of healthcare utilization (17-19). More of these resources are required to enable high quality CHD research that can address the need to understand the long-term outcomes and life course for people living with CHD.

Linking and enabling access to similar resources in Australia will provide a significant contribution to these efforts. The Australian and New Zealand Congenital Heart Registry (ANZCHD Registry) is a comprehensive and contemporary data collection of children and adults with CHD across these two countries (20). At the time of writing, there are over 71,000 unique CHD patients in the Australian part of this Registry. Australia also has a comprehensive public health system supported by large, long-term datasets and well-established data linkage institutions (including records of deaths, medical encounters, hospital admissions, drug prescriptions and emergency department encounters, amongst others)(21). We undertook linkage of the Australian patients in the Registry to Australian administrative health records to develop a comprehensive research infrastructure that will drive continued understanding CHD. Here, we outline the methods for dataset preparation, detail linkage processes and provide an initial overview of the data.

## Methods

### Ethics Statement

Australian Human Research Ethics Approval was obtained from the Sydney Local Health District Ethics Review Committee (RPAH Zone) (2019/ETH07472). Data linkage approvals were obtained from the Australian Institute of Health and Welfare Ethics Committee (EO2023/3/1327), the New South Wales Population and Health Services Research Ethics Committee (2022/ETH01341), and the Northern Territory Health and Menzies School of Health Research Human Research Ethics Committee (2023-4518).

### Data Overview: Cohorts and Datasets

The linkage data is comprised of two cohorts. “Cohort 1” includes people with CHD, identified from the Registry and collected from the major CHD specialist centers across Australia and New Zealand (pediatric and adult, as described in Reference 20). “Cohort 2” includes people who were not already identified in Cohort 1 with at least one episode of care that includes a diagnosis of CHD within one of the “admitted patients” datasets, described in Table 1. Eligible diagnoses include codes Q20.0 to Q26.9 from the 10^th^ revision of the International Classification of Disease – Australian Modification (ICD10-AM).

**Table 1.** Overview and description of the datasets included in the linkage between the ANZCHD Registry and Australian administrative health data. EPCC – European Pediatric Cardiology Code.

| Source | Dataset | Jurisdiction | Description | Date Range |
| --- | --- | --- | --- | --- |
| Australia and New Zealand Congenital Heart Disease Registry (ANZCHD Registry) | Patient Table | Australia excluding Western Australia | Person-level demographics for ANZCHD Registry participants. | Jan 2010 – Jun 2023 |
|  | Diagnosis Table | Australia excluding Western Australia | EPCC-coded diagnosis codes for ANZCHD Registry participants. | Jan 2010 – Jun 2023 |
|  | Procedure Table | Australia excluding Western Australia | EPCC-coded procedure codes for ANZCHD Registry participants. | Jan 2010 – Jun 2023 |
| Australian Institute of Health and Welfare | Medicare Benefits Schedule (MBS) | All Australia | Record of claims for medical services that are reimbursable under Medicare, Australia's public health insurance program. | Jan 2010 – Sep 2024 |
|  | Pharmaceutical Benefits Schedule (PBS) | All Australia | Dispensing data for Australia's national drug subsidy program. | Jan 2010 – Sep 2024 |
|  | National Death Index (NDI) | All Australia | National Register for all death's in Australia. | Jan 1984 – Sep 2024 |
|  | National Hospital Morbidity Database (NHMD) | Australia excluding Western Australia and Northern Territory | A federally held resource, compiled of data supplied by state and territory health authorities, a collection of episodes of care from admitted patients in public and private hospitals. | Feb 2009 – Jun 2022 |
|  | National Non-Admitted Patient Emergency Department Care Database (NNAPEDCD) | Australia excluding Western Australia and Northern Territory | A federally held resource, compiled of data supplied by state and territory health authorities, a collection of records for non-admitted patients treated in public hospital emergency departments. | June 2010 – June 2022 |
| Centre for Victorian Data Linkage | Victorian Admitted Episode Dataset (VAED) | Victoria only | A collection of episodes of care from admitted patients in public and private hospitals in Victoria, with detailed cost data included from the Victorian Cost Data Collection. | April 2009 – May 2024 |
|  | Victorian Emergency Minimum Dataset (VEMD) | Victoria only | A collection of records for non-admitted patients treated in public hospital emergency departments in Victoria. | Dec 2009 – May 2024 |
| South Australia – Northern Territory Datalink | Northern Territory Inpatient Activity (NTIA) | Northern Territory only | A collection of episodes of care from admitted patients in public and private hospitals in the Northern Territory. | July 2009 – May 2023 |
|  | Northern Territory Emergency Department Activity Collection (NTEDAC) | Northern Territory only | A collection of records for non-admitted patients treated in public hospital emergency departments in the Northern Territory. | Jan 2010 – June 2022 |

Data from eligible individuals were extracted from administrative health datasets by three different government linkage agencies in Australia (as in Table 1). Federally, the Australian Institute of Health and Welfare (AIHW) holds data for reimbursable claims for medical services under the Medicare Benefits Schedule (MBS), dispensing data for subsidized medications under the Pharmaceutical Benefits Scheme (PBS), and the National Death Index (NDI). The AIHW also holds two collections of state-level data across all Australian States and Territories except the Northern Territory and Western Australia. These include admitted patients’ data from public and private hospitals in the National Hospital Morbidity Database (NHMD), and emergency department presentations data in the National Non-Admitted Patient Emergency Department Care Database (NNAPEDCD).

Data from the Northern Territory was also provided by South Australia-Northern Territory Datalink (SA-NT Datalink). These included admitted patients from public and private hospitals, Northern Territory Inpatient Activity (NTIA), and emergency department presentations, Northern Territory Emergency Department Activity Collection (NTEDAC). Detailed cost data for inpatient admissions and emergency department presentations is uniquely available in Victoria; thus specific linkages with the Centre for Victorian Record Linkage (CVDL) were sought. These were hospital admitted patient data, the Victorian Admitted Episode Dataset (VAED), and emergency presentation data, the Victorian Emergency Department Activity Collection (VEMD). Unfortunately, state-specific privacy legislation and delays in Western Australia data linkage projects prevented the inclusion of Western Australian datasets in this project (22).

Finally, data from the Registry were also included for people in Cohort 1. This includes 3 tables, a “Patient” table outlining peoples’ key demographic information, and “Diagnosis” and “Procedure” Tables that included diagnosis and procedure codes using the European Pediatric Cardiology Code (EPCC) (23).

### Data Collection and Linkage

For Cohort 1, data was extracted directly from the Registry. The specific data collection and eligibility criteria for the Registry have been previously described (20). A linkage file of approved person identifiers was prepared, and records with missing data in key identifying variables (name and date of birth) were excluded. A prepared, cleaned dataset of Registry patients with person identifiers for linkage was provided to the accredited linkage agencies outlined above to complete probabilistic linkage to the requested administrative health datasets.

For Cohort 2, all diagnosis codes from to an episode of care in the hospital discharge data were checked against the selection criteria. Once the individuals who met these criteria were identified, their records from the requested datasets were extracted. To be able to identify when the same individual in Cohort 2 was selected across jurisdictions and data must be shared between institutions and duplicates identified. This process is still ongoing between the AIHW and CVDL but has not been completed at the time of publication. Furthermore, local legislation in the Northern Territory does not allow sharing of these details at all, and duplicates will not be able to be identified for that portion of cohort 2.

### Data Cleaning and Curation

Since these linkage processes utilize probabilistic methods, the data were reviewed carefully, to identify and exclude potential incorrect linkages. Details of these processes are included in the Data Supplement.

To provide clear definitions of the dataset and to ensure analysis is conducted on consistent groups within this dataset, groupings were identified in each cohort. For Cohort 1, different patient groups were defined in the same way as outlined in the main data collection for the Registry (20). Briefly, Cohort 1 contains three groups. The main study population includes people with diagnosis of a structural CHD. The transient CHD group includes people who have fetal cardiac structures that have been recorded in the neonatal period but not recorded thereafter, without requiring surgical intervention. This includes diagnoses such as patent foramen ovale and persistent arterial duct. The final miscellaneous group include other conditions that may affect the heart at birth but do not present as a structural defect, such as a cardiomyopathy or valve regurgitations with no structural component recorded.

For Cohort 2, certain ICD-10AM codes within the selection criteria were identified as outside the main structural CHD study population. When occurring in isolation, patent foramen ovale (Q21.11), persistent arterial duct with no repair and no follow up after 12 months (Q25.0), and unspecified CHD (Q24.9 or Q24.89), were excluded from the structural CHD cohort.

### Data Analysis

Data preparation and analysis was completed in SAS v9.4 and R Statistical Software v4.3.2. Patients were categorized by CHD complexity using an automated algorithm that reads EPCC codes and applies the complexity rules outlined in the 2020 ESC guidelines for the management of adult CHD (24) (25, 26). For Cohort 2, EPCC codes were not available since these records did not have a link to the Registry coding. In these cases, ICD10-AM codes were translated to EPCC codes to then apply the CHD complexity algorithm. Code translation utilized methods that have been developed for the Registry’s primary data collection (23). Age was calculated at the end of the data collection period, September 2024 (or age at death). An independent samples t-test was used to compare mean age between cohorts. We assessed the association between cohort and disease complexity using the Chi-squared test of independence, reporting Cramér’s V as a measure of effect size.

## Results

For Cohort 1, there were 82,998 patient records identified from the registry, and after excluding patients from Western Australia or those missing key demographic data, there were 72,681 individuals selected. The linkage process identified 1,261 duplicates, leaving 71,420 individuals in the linkage file. There were 1,235 excluded in the data cleaning process, with 70,185 individuals in the final linked Cohort 1 dataset. Of these 58,523 were identified as having structural CHD: the main study population (Figure 1).

**Figure 1.**
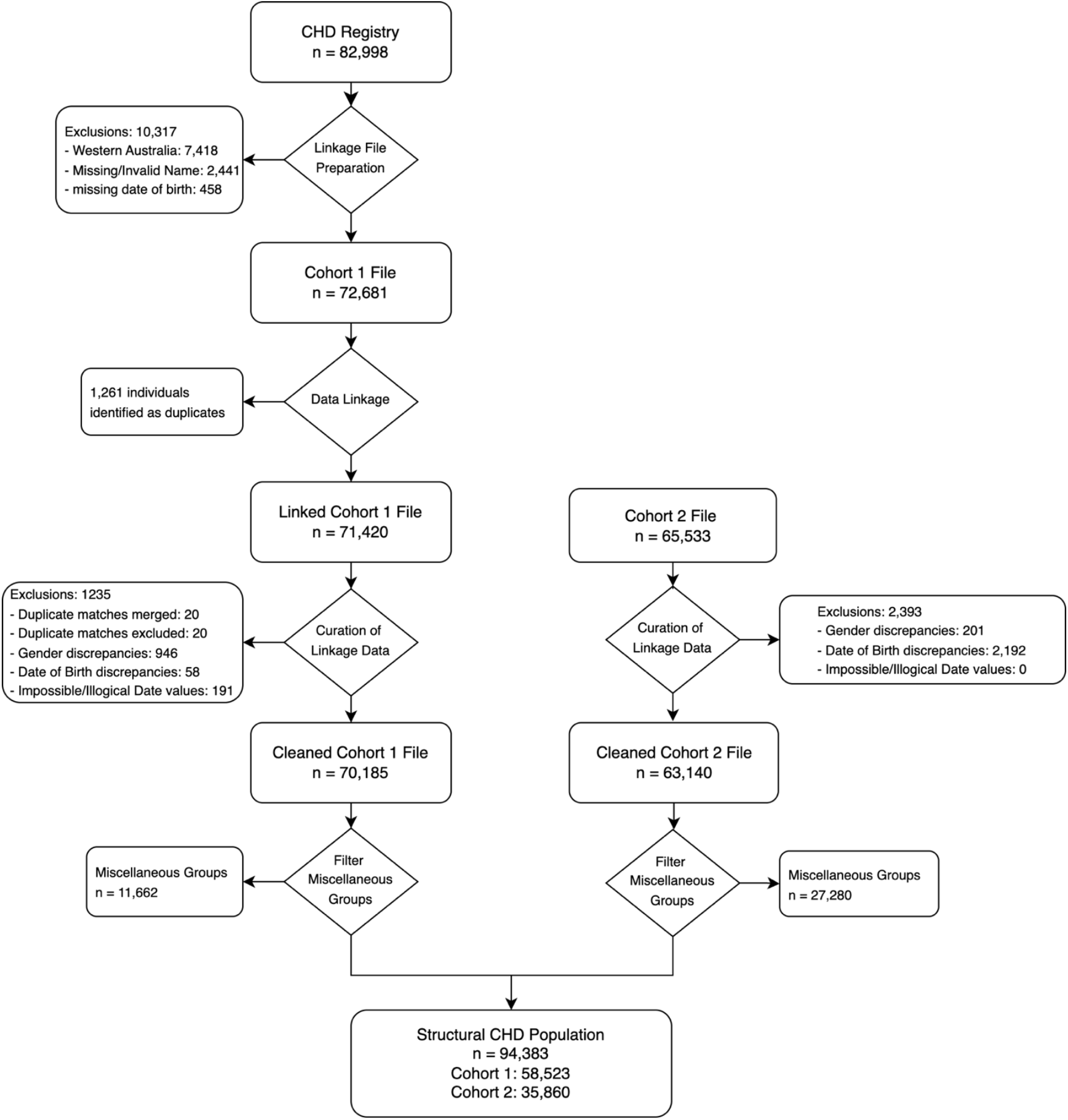
Data collection flowchart for the linked dataset.

Individuals eligible for Cohort 2 were identified by the data linkage agencies. The AIHW identified 40,097 people in the NHMD, CVDL identified 24,929 people in the VAED, and SA-NT Datalink identified 507 people in the NTIA. Data cleaning occurred in the same way as Cohort 1, excluding 2,393 individuals and leaving a cleaned dataset with 63,140 individuals. Finally, there were 35,860 individuals identified with structural CHD and selected as part of the main study population (Figure 1).

Of the 94,383 individuals in the structural CHD population, the median age was 25 years (IQR: 11-43) and 48% were females. There were 6,883 (7%) deaths and there were 32% with mild CHD, 31% with moderate CHD, and 15% with Severe CHD. The median age for Cohorts 1 and 2 was 23 years and 36 years, respectively. On average, Cohort 1 was 14 years younger than Cohort 2 (95% CI: 13.2 – 13.9, p<0.001). A Chi-square test of independence revealed a significant association between CHD complexity and the cohorts (χ2 (df=3, N=94,383) = 7433.1, p<0.001). Cohort 1 had a higher proportion of severe CHD lesions (20%) compared to Cohort 2 (6%), representing a moderate effect (Cramér’s V = 0.28) (Table 2). The miscellaneous groups that were excluded from the main structural CHD population are outlined in Table 3.

**Table 2.**
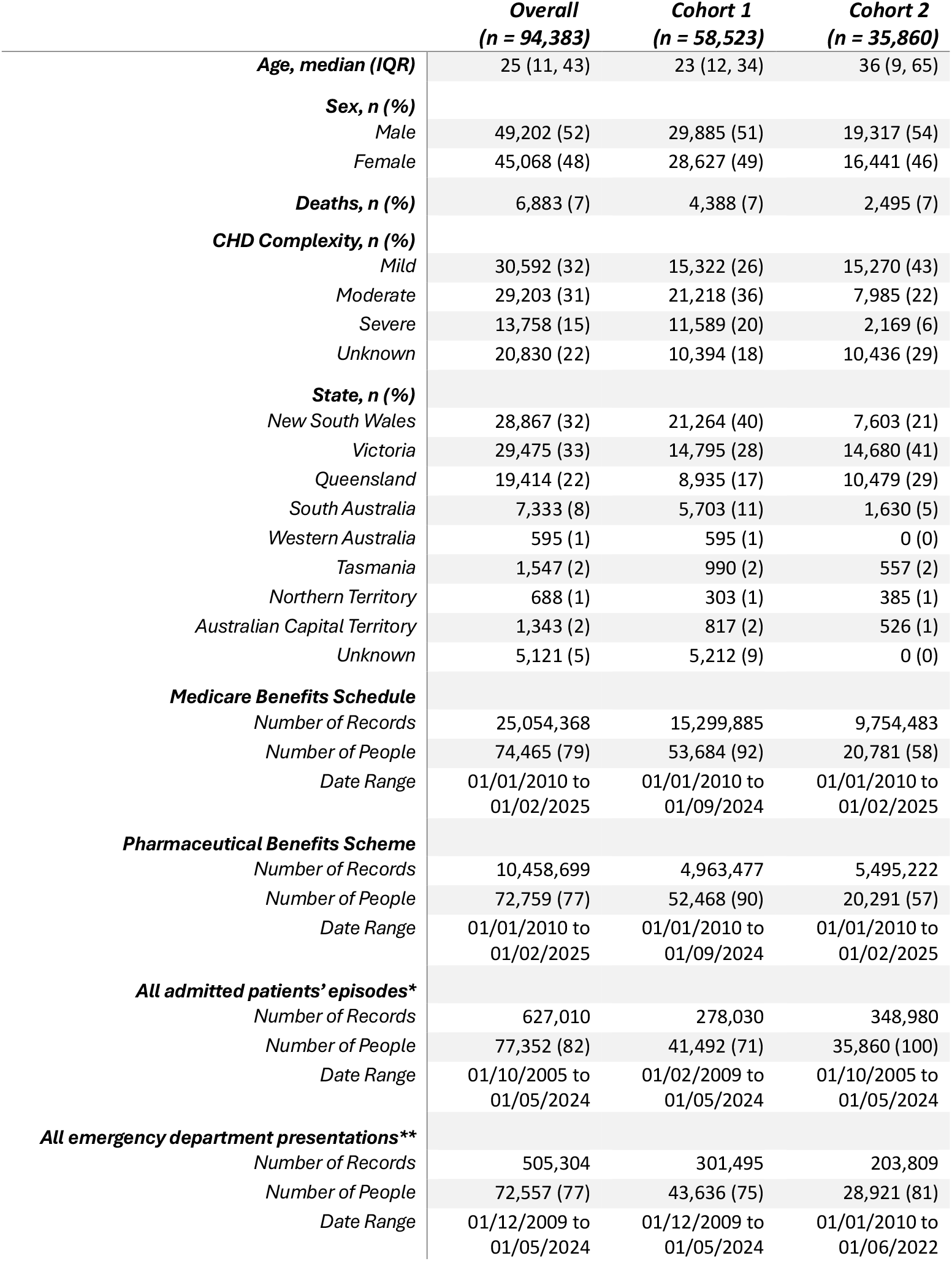
Cohort Description for the full linked dataset of structural CHD patients, including demographic details and the number of linked healthcare encounter records in each dataset. * “All admitted patients’ episodes” combines records from the National Hospita l Morbidity Database, Victorian Admitted Episode Dataset, and the Northern Territory Inpatient Activity tables. ** “All emergency department presentations” combines records from the National Non-Admitted Patient Emergency Department Care Database, Victorian Emergency Minimum Dataset, and the Northern Territory Emergency Department Activity Collection tables.

**Table 3.**
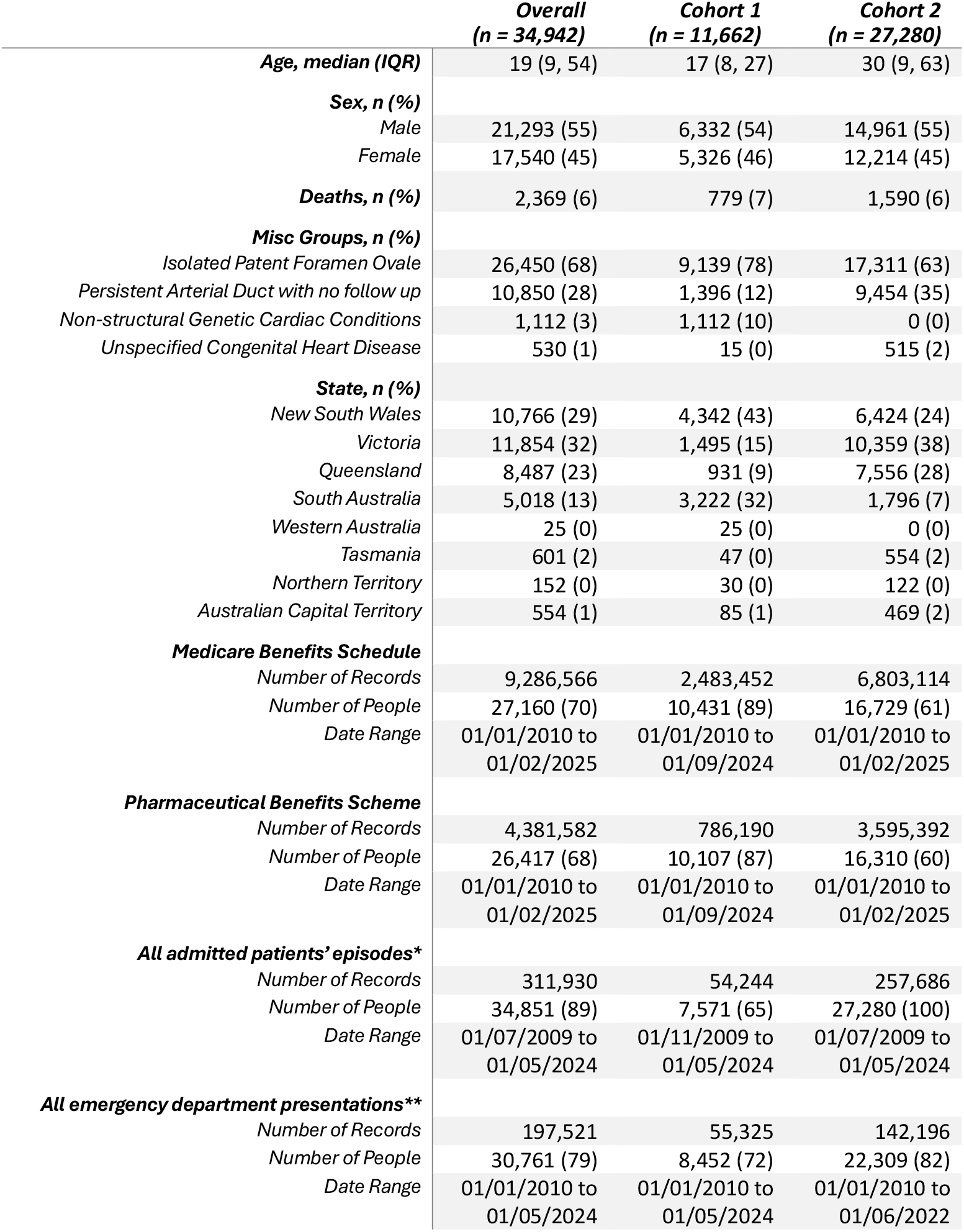
Cohort Description for the full linked dataset of miscellaneous groups, including demographic details and the number of linked healthcare encounter records in each dataset. * “All admitted patients’ episodes” combines records from the National Hospital M Morbidity Database, Victorian Admitted Episode Dataset, and the Northern Territory Inpatient Activity tables. ** “All emergency department presentations” combines records from the National Non-Admitted Patient Emergency Department Care Database, Victorian Emergency Minimum Dataset, and the Northern Territory Emergency Department Activity Collection tables.

|  | **Overall**<br>**(n = 34,942)** | **Cohort 1**<br>**(n = 11,662)** | **Cohort 2**<br>**(n = 27,280)** |
| --- | --- | --- | --- |
| **Age, median (IQR)** | 19 (9, 54) | 17 (8, 27) | 30 (9, 63) |
| **Sex, n (%)** |  |  |  |
| Male | 21,293 (55) | 6,332 (54) | 14,961 (55) |
| Female | 17,540 (45) | 5,326 (46) | 12,214 (45) |
| **Deaths, n (%)** | 2,369 (6) | 779 (7) | 1,590 (6) |
| **Misc Groups, n (%)** |  |  |  |
| Isolated Patent Foramen Ovale | 26,450 (68) | 9,139 (78) | 17,311 (63) |
| Persistent Arterial Duct with no follow up | 10,850 (28) | 1,396 (12) | 9,454 (35) |
| Non-structural Genetic Cardiac Conditions | 1,112 (3) | 1,112 (10) | 0 (0) |
| Unspecified Congenital Heart Disease | 530 (1) | 15 (0) | 515 (2) |
| **State, n (%)** |  |  |  |
| New South Wales | 10,766 (29) | 4,342 (43) | 6,424 (24) |
| Victoria | 11,854 (32) | 1,495 (15) | 10,359 (38) |
| Queensland | 8,487 (23) | 931 (9) | 7,556 (28) |
| South Australia | 5,018 (13) | 3,222 (32) | 1,796 (7) |
| Western Australia | 25 (0) | 25 (0) | 0 (0) |
| Tasmania | 601 (2) | 47 (0) | 554 (2) |
| Northern Territory | 152 (0) | 30 (0) | 122 (0) |
| Australian Capital Territory | 554 (1) | 85 (1) | 469 (2) |
| **Medicare Benefits Schedule** |  |  |  |
| Number of Records | 9,286,566 | 2,483,452 | 6,803,114 |
| Number of People | 27,160 (70) | 10,431 (89) | 16,729 (61) |
| Date Range | 01/01/2010 to<br>01/02/2025 | 01/01/2010 to<br>01/09/2024 | 01/01/2010 to<br>01/02/2025 |
| **Pharmaceutical Benefits Scheme** |  |  |  |
| Number of Records | 4,381,582 | 786,190 | 3,595,392 |
| Number of People | 26,417 (68) | 10,107 (87) | 16,310 (60) |
| Date Range | 01/01/2010 to<br>01/02/2025 | 01/01/2010 to<br>01/09/2024 | 01/01/2010 to<br>01/02/2025 |
| **All admitted patients' episodes\*** |  |  |  |
| Number of Records | 311,930 | 54,244 | 257,686 |
| Number of People | 34,851 (89) | 7,571 (65) | 27,280 (100) |
| Date Range | 01/07/2009 to<br>01/05/2024 | 01/11/2009 to<br>01/05/2024 | 01/07/2009 to<br>01/05/2024 |
| **All emergency department presentations\*\*** |  |  |  |
| Number of Records | 197,521 | 55,325 | 142,196 |
| Number of People | 30,761 (79) | 8,452 (72) | 22,309 (82) |
| Date Range | 01/01/2010 to<br>01/05/2024 | 01/01/2010 to<br>01/05/2024 | 01/01/2010 to<br>01/06/2022 |

The most common CHD lesions in Cohort 1 were Ventricular Septal Defect (14,019, 24%), Atrial Septal Defect (5,698, 10%), Aortic Valve Disorders (4,770, 8%), Coarctation of the Aorta (4,568, 8%), and Persistent Arterial Duct (4,176, 7%). For Cohort 2 the most common lesions were Aortic Valve Disorders (9,213, 25%), Ventricular Septal Defect (7,044, 19%), Atrial Septal Defect (6,527, 18%), Pulmonary Valve Disorders (2,144, 6%), and Persistent Arterial Duct (1,348, 4%) (Figure 2).

**Figure 2.**
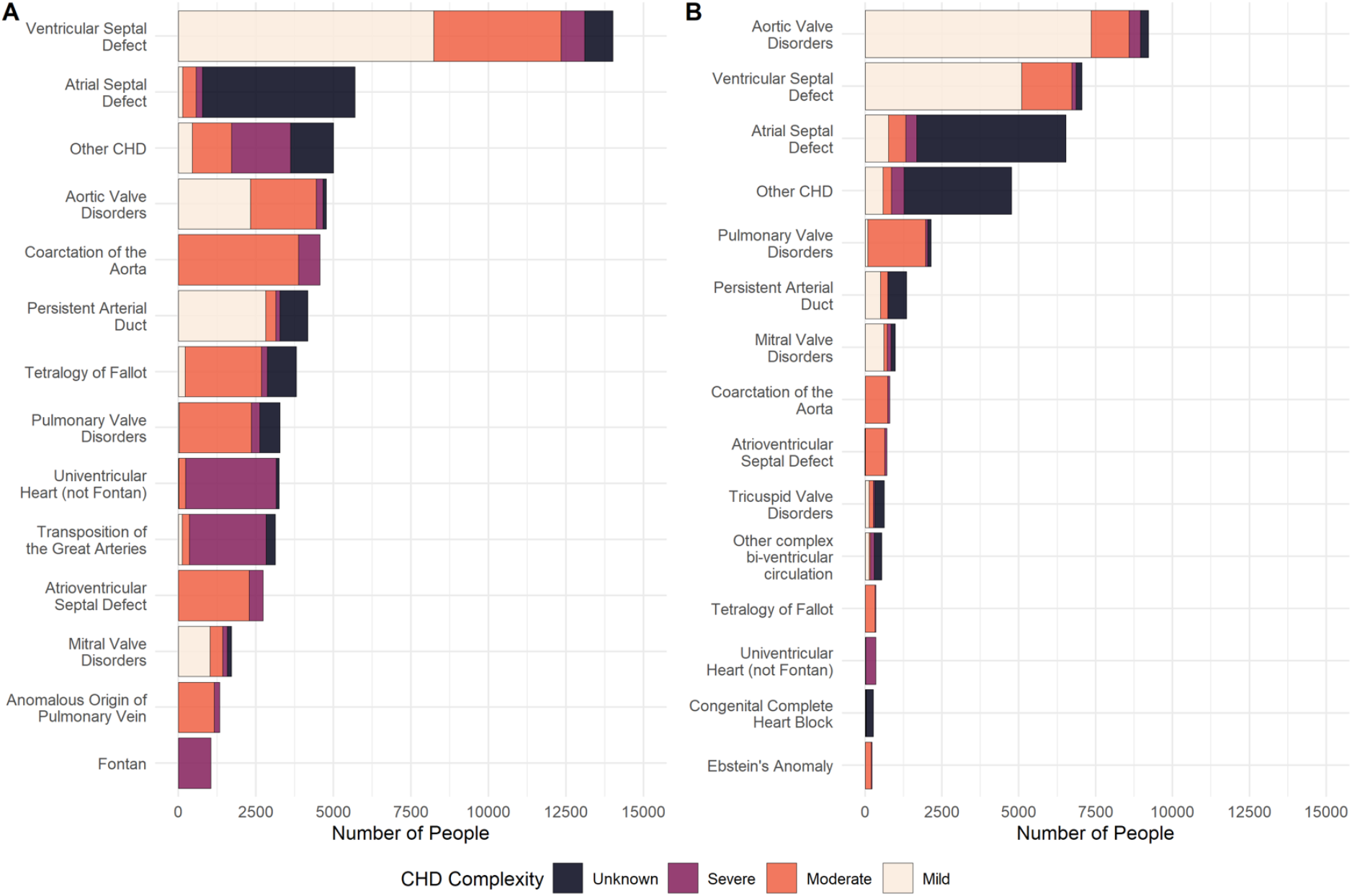
Congenital Heart Disease (CHD) diagnoses in the structural CHD Cohort. The most frequently occurring CHD diagnoses in Cohort 1 (A) and Cohort 2 (B).

## Discussion

The Australia and New Zealand Congenital Heart Disease Registry is one of the world’s largest collections of people with CHD. With a detailed collection of healthcare interactions for over a 10-year period this represents a significant enrichment of the Registry data. We have conducted a comprehensive data curation process to make this data ready for use. This data represents a significant piece of research infrastructure that will drive high quality research, answering the call made by Diller *et. al*. (6) for CHD research to shift its focus to long-term and life course outcomes. The inclusion of Cohort 2 in this dataset provides an opportunity to further improve upon the Registry’s data collection by identifying any individuals who were missed in the primary collection from Australia’s major CHD specialist centers. Interestingly, Cohort 2 tended to be older with more mild CHD lesions, which would be consistent with people who might not be known to specialist CHD service and are potentially lost to follow up.

Currently, linkages to pediatric CHD cohorts in New South Wales have helped understanding of the outcomes and costs associated with cardiac surgery in the first year of life (27), the associations between the age when pediatric cardiac surgery is performed and cardiovascular outcomes (14), and to extrapolate the effect of cardiac surgery in the first year of life to school-aged outcomes (28). These contributions focus on pediatric outcomes covering a single state in Australia. This current project will provide an extension to the existing research by expanding the linkage to a national scale and including follow up of adults.

Administrative data has often utilized mortality data in survival analysis. A 2016 meta-analysis into long-term survival in CHD identified 16 studies, 13 of which accessed administrative records, and seven of these having follow up of 15 years or greater (29). Survival analysis in a congenital disease population requires whole-of-life follow up since “exposure” begins at birth and linkage to administrative data can provide the required long-term data. More recent studies have made use of administrative data to conduct analysis with long-term follow up to 68 years or age (16, 30) in 2017. Our data presented here will provide important contributions to the understanding long-term survival with similar follow up time and more contemporary data.

Dellborg *et. al*. (16) have demonstrated survival in CHD subjects of 75% past 68 years of age, highlighting the importance of research focusing on life course outcomes of people living with CHD. The use of routinely collected administrative data and registry data in 9 studies has allowed for a powerful meta-analysis of 684,200 individuals, demonstrating an increased risk of stroke, heart failure, and coronary artery disease (31). This is an excellent example of how administrative datasets with large population-level data and long follow up times are being leveraged to expand CHD research to focus on the life course.

Administrative health data also provides an excellent tool for undertaking healthcare utilization research, which is especially relevant to the high healthcare needs experienced by the CHD population. A major systematic review of 21 studies showed a general increase in outpatient care and a reduction in hospitalizations but also noted increases in unplanned hospitalization and emergency care (32). Benderly *et. al*. (18) utilized comprehensive electronic medical records to uncovering drivers in healthcare utilization amongst important demographic factors. These examples show how administrative data can be utilized to understand population-level trends in healthcare utilization and associations between healthcare utilization and social determinates of health.

Despite this great potential for use of routinely collected data, these data were not originally collected for research and important limitations must be considered. The linkage process often utilizes probabilistic techniques and linkage errors are a potential cause of bias in this data. This is hard to account for as often the linkage process is obscured from researchers who conduct the analyses, due to important privacy considerations (33). Data linkage in Australia is quite mature with studies reporting error rates of less than 1% (34). The data curation steps outlined in this paper were taken to further assess the linkage results from the perspective of the researcher.

Also, these data only collect information regarding funded interactions, and interactions that are self-funded and claims that are not submitted will be missing (35). This is unusual in Australia, where Medicare provides a “universal” insurance scheme for all children and adults.

Specific to this current project, some legislative limitations exist in achieving National coverage, where Western Australian data has not been included (for state-specific legislative reasons), and identifying duplicates between states has been difficult for cohort (22, 36). Close understanding of the legislative landscape in Australia, and the resulting limitations in the resulting data are important when using these data for observational research. The overview provided in this publication will help future researchers to navigate these challenges.

The accuracy of diagnostic and procedure coding is also crucial, especially in relation to this data where hospital diagnosis coding is used to identify CHD patients. Recent assessment of the coding of congenital abnormalities compared Australia hospital coding against registry coding as a gold standard and found >93% agreement for cardiac abnormalities, suggesting this as a suitable method of cohort selection (37). Finally, understanding the ability for administrative data to accurately identify Indigenous individuals is important. Australian data relies on self-reporting and under-reporting is a recognized issue for these data (38).

This linkage project of the ANZCHD Registry and Australian administrative health data represents the union of two world class databases that will contribute to the challenges of improving CHD outcomes. The collection methodology and data curation outlined here aims to provide this dataset as research infrastructure, an accessible and reliable tool for ongoing contemporary CHD research. Future enrichment and improvement of this linked data will be sought to maintain this data as an ongoing, relevant resource for future research.

## Acknowledgements

The Congenital Heart Alliance of Australia and New Zealand consist of key academic and clinical members of the CHD community across Australia and New Zealand. The data that are described in this manuscript, and that makes up the Australian National CHD Registry, were provided by the clinical teams in these groups. Its members include; Rachael Cordina MBBS, PhD^a,b,c^, Julian Ayer BSc, MMBS, PhD^a,d^, Michael Cheung MD, PhD^e^, Leeanne Grigg MBBS, FRACP^f^, Robert Justo MBBS, FRACP^g^, Ryan Maxwell MBBS, FRACP^h^, Gavin Wheaton MBBS,FRACP^i^, Patrick Disney MBBS^j^, Deane Yim MBChB, FRACP^k,l^, and Clare O’Donnell MBChB SM, FRACP^m^

^a^ University of Sydney, Camperdown, NSW, 2050, Australia

^b^ Heart Research Institute, 7 Eliza St, Newtown NSW 2042, Australia

^c^ Royal Prince Alfred Hospital, 50 Missenden Rd, Camperdown NSW 2050, Australia

^d^ Sydney Children’s Hospital Network, Corner Hawkesbury Road and, Hainsworth St, Westmead NSW 2145, Australia

^e^ The Royal Children’s Hospital, 50 Flemington Rd, Parkville VIC 3052, Australia

^f^ The Royal Melbourne Hospital, 300 Grattan St, Parkville VIC 3052, Australia

^g^ Queensland Children’s Hospital, 501 Stanley St, South Brisbane QLD 4101, Australia

^h^ The Prince Charles Hospital, 627 Rode Rd, Chermside QLD 4032, Australia

^i^ Women’s and Children’s Hospital, 72 King William Rd, North Adelaide SA 5006, Australia

^j^ Royal Adelaide Hospital, Port Rd, Adelaide SA 5000, Australia

^k^ Perth Children’s Hospital, 15 Hospital Ave, Nedlands WA 6009, Australia

^l^ Sir Charles Gairdner Hospital, Hospital Ave, Nedlands WA 6009, Australia

^m^ Green Lane Paediatric and Congenital Cardiac Service, Starship Children’s Hospital, Victoria Street West, Auckland 1142, New Zealand

## Author Contributions

All authors contributed to study conceptualizations. Calum Nicholson performed the protocol development, data preparation, data visualization, and manuscript drafting. Geoff Strange contributed to protocol development, data preparation and manuscript editing. William Baxter contributed to data preparation and manuscript editing. Larrissa Lloyd contributed to protocol development and manuscript editing. David Celemajer contributed to protocol development, data preparation, and manuscript editing.

## Conflict of Interest

None Declared

## Funding

This work was supported by the Department of Health and Ageing, Australian Government through an Accelerated Research Grant – congenital heart disease stream from the Medical Research Futures Fund. The

grants name is “An Australian Study of the Outcomes and Burden of Congenital Heart Disease” [grant number ARGCHDG000028]”.

## Data Availability

Due to the sensitive and private nature of the health information held in the Registry, it is not publicly available. Deidentified access can be sought by contributors to the Registry, enquires can be made at https://www.chaanz.org.au/.

## Notes

### Competing Interest Statement

The authors have declared no competing interest.

### Clinical Trial

Not a clinical trial

## References

1. Erikssen G, Liestøl K, Seem E, et al. Achievements in congenital heart defect surgery: a prospective, 40-year study of 7038 patients. Circulation. 2015;131(4):337–46; discussion 46.

2. Grown-up congenital heart (GUCH) disease: current needs and provision of service for adolescents and adults with congenital heart disease in the UK. Heart. 2002;88 Suppl 1(Suppl 1):i1–14.

3. Abdurrahman L. Adult congenital heart disease update. Current Problems in Pediatric and Adolescent Health Care. 2023:101399.

4. Nicolae M, Gentles T, Strange G, et al. Adult Congenital Heart Disease in Australia and New Zealand: A Call for Optimal Care. Heart, Lung and Circulation. 2019;28(4):521–9.

5. Liu A, Diller GP, Moons P, Daniels CJ, Jenkins KJ, Marelli A. Changing epidemiology of congenital heart disease: effect on outcomes and quality of care in adults. Nat Rev Cardiol. 2023;20(2):126–37.

6. Diller GP, Arvanitaki A, Opotowsky AR, et al. Lifespan Perspective on Congenital Heart Disease Research: JACC State-of-the-Art Review. J Am Coll Cardiol. 2021;77(17):2219–35.

7. van der Linde D, Konings EE, Slager MA, et al. Birth prevalence of congenital heart disease worldwide: a systematic review and meta-analysis. J Am Coll Cardiol. 2011;58(21):2241–7.

8. van der Velde ET, Vriend JW, Mannens MM, Uiterwaal CS, Brand R, Mulder BJ. CONCOR, an initiative towards a national registry and DNA-bank of patients with congenital heart disease in the Netherlands: rationale, design, and first results. Eur J Epidemiol. 2005;20(6):549–57.

9. Tobler D, Schwerzmann M, Bouchardy J, et al. Swiss Adult Congenital HEart disease Registry (SACHER) - rationale, design and first results. Swiss Med Wkly. 2017;147:w14519.

10. Ombelet F, Goossens E, Willems R, et al. Creating the BELgian COngenital heart disease database combining administrative and clinical data (BELCODAC): Rationale, design and methodology. Int J Cardiol. 2020;316:72–8.

11. Watelle L, Louis-Olivier R, Jonathan L-S, et al. The Quebec Congenital Heart Disease Registry: A Model of Prospective Databank to Facilitate Research in Congenital Cardiology. CJC Pediatric and Congenital Heart Disease. 2023.

12. Christopher MS, Christopher Martin S, Li-Ching C, et al. Leveraging electronic health records for data science: common pitfalls and how to avoid them. The Lancet Digital Health. 2022.

13. Casey JA, Schwartz BS, Stewart WF, Adler NE. Using Electronic Health Records for Population Health Research: A Review of Methods and Applications. Annual Review of Public Health. 2016;37(Volume 37, 2016):61–81.

14. Lain SJ, Blue GM, O’Malley BR, et al. Using novel data linkage of congenital heart disease biobank data with administrative health data to identify cardiovascular outcomes to inform genomic analysis. Int J Popul Data Sci. 2023;8(1):2150.

15. Downing KF, Nembhard WN, Rose CE, et al. Survival From Birth Until Young Adulthood Among Individuals With Congenital Heart Defects: CH STRONG. Circulation. 2023;148(7):575–88.

16. Dellborg M, Giang KW, Eriksson P, et al. Adults With Congenital Heart Disease: Trends in Event-Free Survival Past Middle Age. Circulation. 2023;147(12):930–8.

17. Islam S, Kaul P, Tran DT, Mackie AS. Health Care Resource Utilization Among Children With Congenital Heart Disease: A Population-Based Study. The Canadian journal of cardiology. 2018;34(10):1289–97.

18. Benderly M, Buber J, Kalter-Leibovici O, et al. Health Service Utilization Patterns Among Adults With Congenital Heart Disease: A Population-Based Study. J Am Heart Assoc. 2021;10(2):e018037.

19. Edelson JB, Rossano JW, Griffis H, et al. Emergency Department Visits by Children With Congenital Heart Disease. J Am Coll Cardiol. 2018;72(15):1817–25.

20. Nicholson C, Strange G, Ayer J, et al. A national Australian Congenital Heart Disease registry; methods and initial results. International Journal of Cardiology Congenital Heart Disease. 2024;17:100538.

21. Smith M, Flack F. Data linkage in Australia: The first 50 years. International Journal of Environmental Research and Public Health. 2021;18(21):11339.

22. Lloyd LK, Nicholson C, Strange G, Celermajer DS. State- and territory-based differences that impede the establishment of a harmonised national registry. Aust Health Rev. 2025;49.

23. Chami J, Strange G, Nicholson C, Celermajer DS. Towards a Unified Coding System for Congenital Heart Diseases. Circ Cardiovasc Qual Outcomes. 2021;14(7):e008216.

24. Baumgartner H, De Backer J, Babu-Narayan SV, et al. 2020 ESC Guidelines for the management of adult congenital heart disease: The Task Force for the management of adult congenital heart disease of the European Society of Cardiology (ESC). Endorsed by: Association for European Paediatric and Congenital Cardiology (AEPC), International Society for Adult Congenital Heart Disease (ISACHD). European Heart Journal. 2020;42(6):563–645.

25. Chami J, Nicholson C, Baker D, Cordina R, Strange G, Celermajer DS. Improved complexity stratification in congenital heart disease; the impact of including procedural data on accuracy and reliability. International Journal of Cardiology Congenital Heart Disease. 2024;16:100510.

26. Chami J, Strange G, Baker D, et al. Algorithmic complexity stratification for congenital heart disease patients. International Journal of Cardiology Congenital Heart Disease. 2023;11:100430.

27. Claire ML, Claire ML, Samantha JL, et al. Mortality, rehospitalizations and costs in children undergoing a cardiac procedure in their first year of life in New South Wales, Australia. International Journal of Cardiology. 2017.

28. Claire ML, Claire ML, David SW, et al. School-Age Developmental and Educational Outcomes Following Cardiac Procedures in the First Year of Life: A Population-Based Record Linkage Study. Pediatric Cardiology. 2019.

29. Best KE, Rankin J. Long-Term Survival of Individuals Born With Congenital Heart Disease: A Systematic Review and Meta-Analysis. J Am Heart Assoc. 2016;5(6).

30. Dellborg MD, Giang WGK, Eriksson PE, et al. Long-term survival in adults with congenital heart disease: a nationwide, register-based cohort study. European Heart Journal. 2022;43(Supplement_2).

31. Wang T, Chen L, Yang T, et al. Congenital Heart Disease and Risk of Cardiovascular Disease: A Meta-Analysis of Cohort Studies. Journal of the American Heart Association. 2019;8(10):e012030.

32. Willems R, Werbrouck A, De Backer J, Annemans L. Real-world healthcare utilization in adult congenital heart disease: a systematic review of trends and ratios. Cardiol Young. 2019;29(5):553–63.

33. Harron K, Dibben C, Boyd J, et al. Challenges in administrative data linkage for research. Big Data & Society. 2017;4(2):2053951717745678.

34. Boyd JH, Randall SM, Ferrante AM, et al. Accuracy and completeness of patient pathways – the benefits of national data linkage in Australia. BMC Health Services Research. 2015;15(1):312.

35. Harbaugh CM, Cooper JN. Administrative databases. Seminars in Pediatric Surgery. 2018;27(6):353–60.

36. Lloyd LK, Nicholson C, Strange G, Celermajer DS. The burdensome logistics of data linkage in Australia - the example of a national registry for congenital heart disease. Aust Health Rev. 2024;48(1):8–15.

37. Schneuer FJ, Lain SJ, Bell JC, Goldsmith S, McIntyre S, Nassar N. The accuracy of hospital discharge data in recording major congenital anomalies in Australia. Birth Defects Res. 2021;113(18):1313–23.

38. Thompson SC, Woods JA, Katzenellenbogen JM. The quality of Indigenous identification in administrative health data in Australia: insights from studies using data linkage. BMC Medical Informatics and Decision Making. 2012;12(1):133.

